# Variant electrical activation and recovery in normal human hearts revealed by noninvasive electrocardiographic imaging

**DOI:** 10.1101/2024.04.29.24306428

**Authors:** Job Stoks, Kiran Patel, Bianca van Rees, Uyen Chau Nguyen, Casper Mihl, Peter M Deissler, Rachel MA ter Bekke, Ralf Peeters, Johan Vijgen, Paul Dendale, Fu Ng, Matthijs JM Cluitmans, Paul GA Volders

**Affiliations:** Department of Cardiology, Cardiovascular Research Institute Maastricht (CARIM), Maastricht University Medical Center+, the Netherlands; Department of Advanced Computing Sciences, Maastricht University, Maastricht, the Netherlands; UHasselt, Faculty of Medicine and Life Sciences, Diepenbeek, Belgium; Department of Cardiology, Hartcentrum, Jessa Hospital, Hasselt, Belgium; National Heart and Lung Institute (NHLI), Imperial College London; Department of Radiology and Nuclear Medicine, Maastricht University Medical Center+, the Netherlands

**Keywords:** ECGI, ventricles, electrophysiology, variability, normal, dynamics

## Abstract

**Background and aims:** Although electrical activity of the normal human heart is well-characterized by the 12-lead electrocardiogram, detailed insights into within-subject and between-subject variations of ventricular activation and recovery by noninvasive electroanatomic mapping are lacking. We characterized human epicardial activation and recovery within and between normal subjects using noninvasive electrocardiographic imaging (ECGI) as a basis to better understand pathology.

**Methods:** Epicardial activation and recovery were assessed by ECGI in 22 normal subjects, 4 subjects with bundle branch block (BBB) and 4 with long-QT syndrome (LQTS). We compared characteristics between the ventricles (LV versus RV), sexes and age groups (<50y/≥50y (years)). Pearson’s correlation coefficient (CC) was used for within-subject and between-subject comparisons.

**Results:** Age of normal subjects averaged 49±14 years, 6/22 were male, and no structural/electrical heart disease was present. Average activation time was longer in LV than in RV, but not different by sex or age. Electrical recovery was similar for the ventricles, but started earlier and was on average shorter in males than females. Median CCs of between-subject comparisons of the ECG signals, activation and recovery patterns were 0.61, 0.32 and 0.19, respectively. Within-subject beat-to-beat comparisons yielded higher CCs (0.98, 0.89 and 0.82, respectively). Activation and/or recovery patterns of patients with BBB or LQTS contrasted significantly with those found in the normal population.

**Conclusion:** Activation and recovery patterns vary profoundly between normal subjects, but are stable individually beat to beat, with a male preponderance to shorter recovery. Individual characterization by ECGI at baseline serves as reference to better understand the emergence, progression and treatment of electrical heart disease.

**Graphical abstract:** 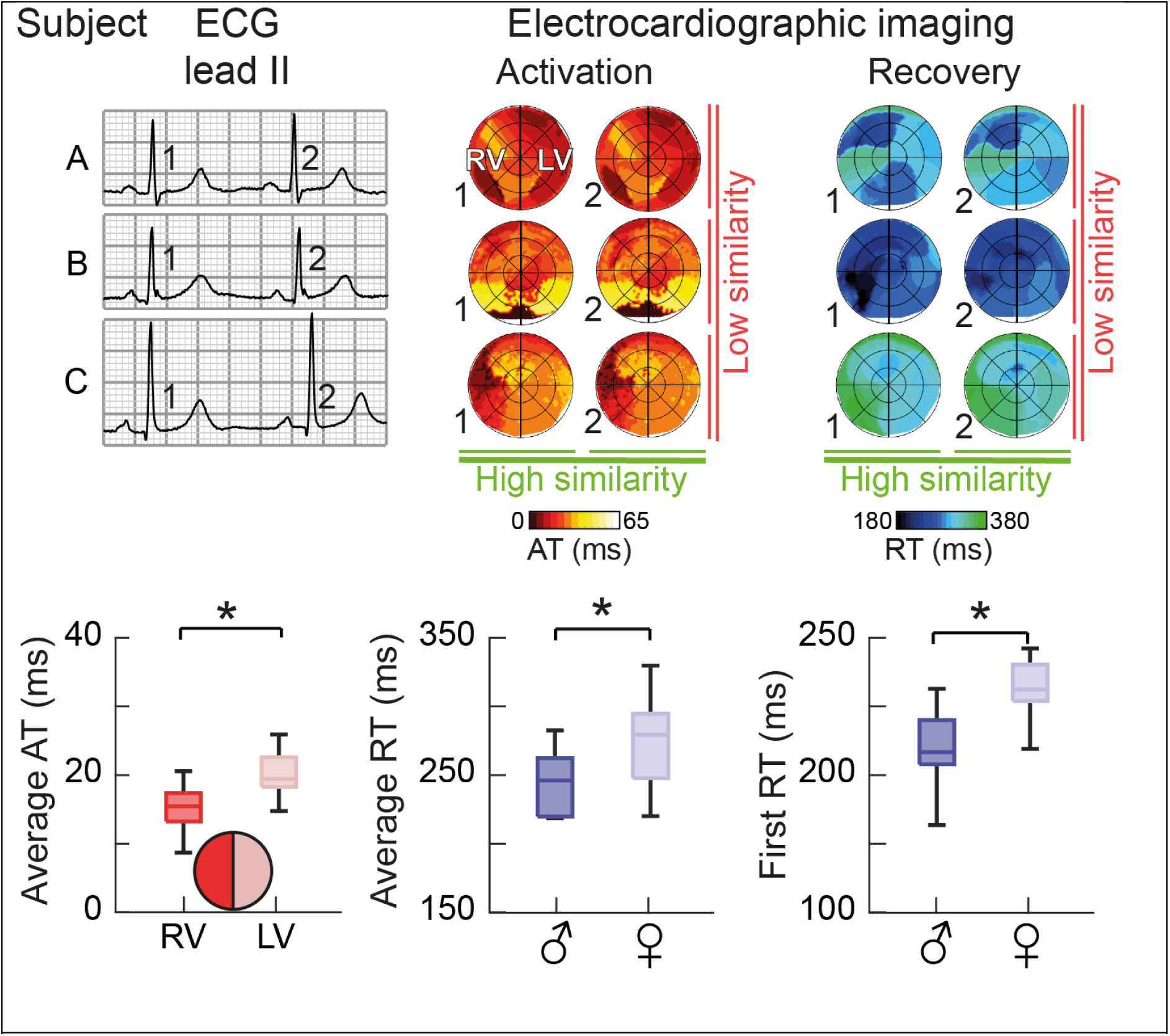
Central illustration. Application of electrocardiographic imaging (ECGI) in healthy controls allows to study normal epicardial activation and recovery patterns. Top: Although all subjects had a normal ECG, we found that their underlying activation time (AT) and recovery time (RT) patterns could be profoundly different. Still, beat-to-beat AT and RT patterns within one subject were relatively similar. Bottom: On a population level, we found that average right-ventricular (RV) AT was lower than left-ventricular (LV) AT, and both first and average RT were lower in males than in females.

## Introduction

Since the introduction of the 3-lead and 12-lead electrocardiogram (ECG), insights into the normal activation and recovery of the human heart have increased dramatically. However, the spatial resolution of the 12-lead ECG limits understanding of localized activation or recovery patterns. Catheter mapping provides high resolution, but is invasive and is only performed in (suspected) disease conditions^1,2^ or on explanted hearts^3–5^.

Electrocardiographic imaging (ECGI) enables noninvasive electrical mapping with a ∼1-cm resolution on the ventricular epicardium^6^. To date, ECGI studies in normal human subjects under physiological conditions are limited to a few only ^7,8^. Detailed insights into the individual characteristics of beat-to-beat dynamics of ECGI-based ventricular electrophysiology and the influence of sex and age are lacking.

We and others have previously validated ECGI (see Supplementary Figure 1). To fill current knowledge gaps, we used it to examine beat-to-beat stability of electrical activation and recovery within and among subjects of different ages, and compared females and males.

Furthermore, to provide a context for electrophysiological variation, we contrasted our results in normal individuals with those of patients with abnormal activation (left and right bundle branch block (LBBB and RBBB)) or recovery patterns (long-QT syndrome (LQTS)).

## Methods

### Study population and consent

This study was approved by the local ethics committees of Maastricht University Medical Center+ (MUMC+), The Netherlands (METC 11-2-043) and the Health Research Authority London, United Kingdom (Surrey; 19/LO/0762), and adhered to the Declaration of Helsinki. All subjects provided written informed consent prior to inclusion. Twenty-two normal human subjects were recruited prospectively from the MUMC+, The Netherlands (n=11) and the Imperial College London, United Kingdom (ICL; n=11). Inclusion criteria for normal subjects were: age ≥18 years, structurally normal heart on echocardiogram, normal 12-lead ECG, and no (suspected) pathology affecting ventricular electrophysiology. Subjects at the MUMC+ underwent a cardiac computed tomography (CT)-scan as part of routine clinical care (often for atypical chest complaints), ruling out significant coronary artery disease or other cardiac abnormalities. Two subjects received metoprolol (50 or 100 mg) for hypertension or atypical complaints. Subjects at ICL were recruited by advertisement, underwent cardiac magnetic resonance imaging (MRI) and were likewise negative for pathological findings on echocardiogram, MRI or ECG.

Eight cardiac patients were recruited by their cardiologist at MUMC+: one with complete RBBB, three with complete LBBB, and four with congenital long-QT syndrome (LQTS). They underwent a cardiac CT-scan as part of the study protocol. All LQTS patients had pathogenic mutations in *KCNQ1, KCNH2* or *SCN5A.* Two patients had previous ventricular tachyarrhythmias and an overtly-prolonged QTc interval (mutations in the *KCHN2* gene (“LQT2 Symp.”) and *SCN5A* gene (“LQT3 Symp.”)), the other two did not (*KCNQ1-* (“LQT1 Asymp.”) and *SCN5A*-gene mutation (“LQT3 Asymp.”)), see also Table 1.

**Table 1:**
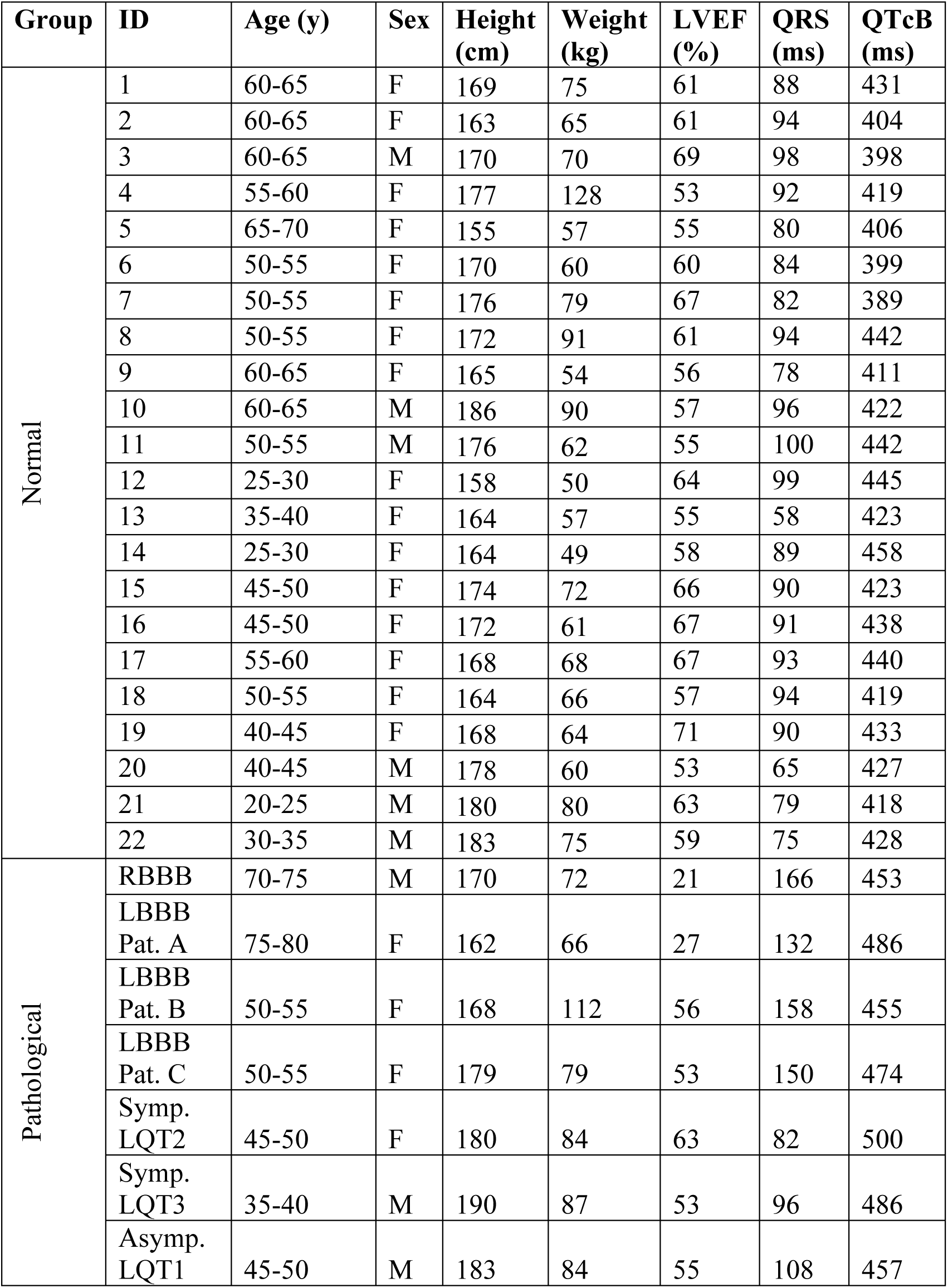
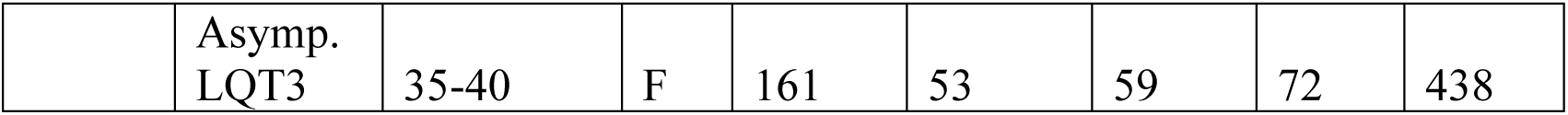
Subject characteristics. LVEF: left-ventricular ejection fraction. SD: standard deviation. QTcB: QT interval corrected according to Bazett’s formula. Pat.: patient. LQTS patients are further divided into symptomatic (“Symp.”; with history of ventricular tachyarrhythmias) and asymptomatic (“Asymp.”; without such history).

| Group | ID | Age (y) | Sex | Height (cm) | Weight (kg) | LVEF (%) | QRS (ms) | QTcB (ms) |
| --- | --- | --- | --- | --- | --- | --- | --- | --- |
| Normal | 1 | 60-65 | F | 169 | 75 | 61 | 88 | 431 |
|  | 2 | 60-65 | F | 163 | 65 | 61 | 94 | 404 |
|  | 3 | 60-65 | M | 170 | 70 | 69 | 98 | 398 |
|  | 4 | 55-60 | F | 177 | 128 | 53 | 92 | 419 |
|  | 5 | 65-70 | F | 155 | 57 | 55 | 80 | 406 |
|  | 6 | 50-55 | F | 170 | 60 | 60 | 84 | 399 |
|  | 7 | 50-55 | F | 176 | 79 | 67 | 82 | 389 |
|  | 8 | 50-55 | F | 172 | 91 | 61 | 94 | 442 |
|  | 9 | 60-65 | F | 165 | 54 | 56 | 78 | 411 |
|  | 10 | 60-65 | M | 186 | 90 | 57 | 96 | 422 |
|  | 11 | 50-55 | M | 176 | 62 | 55 | 100 | 442 |
|  | 12 | 25-30 | F | 158 | 50 | 64 | 99 | 445 |
|  | 13 | 35-40 | F | 164 | 57 | 55 | 58 | 423 |
|  | 14 | 25-30 | F | 164 | 49 | 58 | 89 | 458 |
|  | 15 | 45-50 | F | 174 | 72 | 66 | 90 | 423 |
|  | 16 | 45-50 | F | 172 | 61 | 67 | 91 | 438 |
|  | 17 | 55-60 | F | 168 | 68 | 67 | 93 | 440 |
|  | 18 | 50-55 | F | 164 | 66 | 57 | 94 | 419 |
|  | 19 | 40-45 | F | 168 | 64 | 71 | 90 | 433 |
|  | 20 | 40-45 | M | 178 | 60 | 53 | 65 | 427 |
|  | 21 | 20-25 | M | 180 | 80 | 63 | 79 | 418 |
|  | 22 | 30-35 | M | 183 | 75 | 59 | 75 | 428 |
| Pathological | RBBB | 70-75 | M | 170 | 72 | 21 | 166 | 453 |
|  | LBBB Pat. A | 75-80 | F | 162 | 66 | 27 | 132 | 486 |
|  | LBBB Pat. B | 50-55 | F | 168 | 112 | 56 | 158 | 455 |
|  | LBBB Pat. C | 50-55 | F | 179 | 79 | 53 | 150 | 474 |
|  | Symp. LQT2 | 45-50 | F | 180 | 84 | 63 | 82 | 500 |
|  | Symp. LQT3 | 35-40 | M | 190 | 87 | 53 | 96 | 486 |
|  | Asymp. LQT1 | 45-50 | M | 183 | 84 | 55 | 108 | 457 |
|  | Asymp.<br>LQT3 | 35-40 | F | 161 | 53 | 59 | 72 | 438 |

### Study procedure

The study procedure and ECGI analysis are shown in Figure 1. In both MUMC+ and ICL, ∼200 Ag-AgCl electrodes (ActiveTwo, BioSemi, The Netherlands) were attached to the participant’s torso to record body-surface potentials at a sampling frequency of 2048 Hz for 11±7 minutes at rest (Figure 1A). At MUMC+, subjects then underwent a non-contrast low-dose thoracic CT-scan to image the electrode positions and coronary CT angiography with intravenous administration of iodinated contrast medium to image the heart at end-diastole. Subjects at ICL received a 1.5-T MRI-scan to image the heart geometry at end-diastole and the positions of MRI-safe markers that replaced the body-surface electrodes at identical anatomical locations.

**Figure 1:**
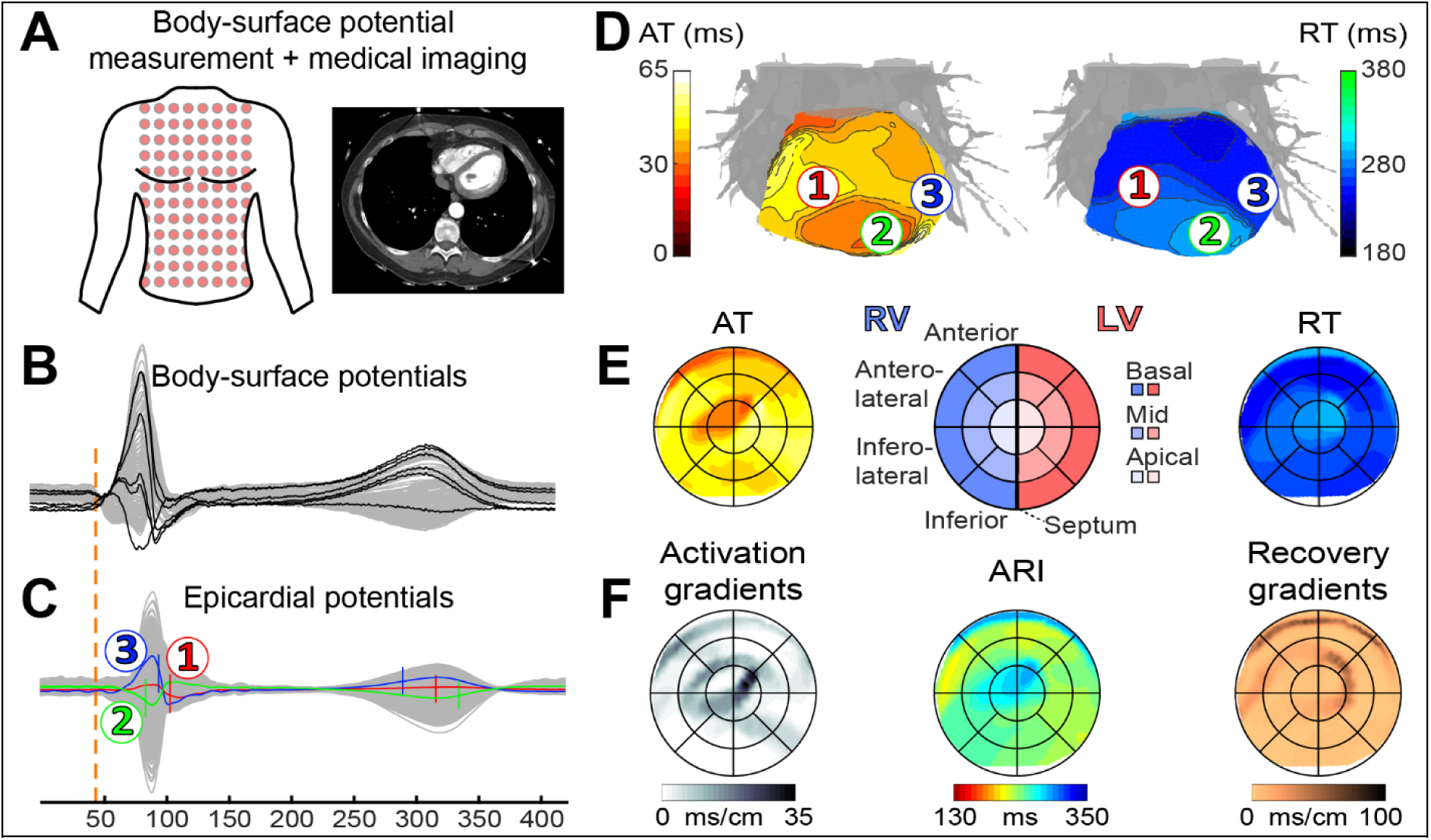
ECGI study procedure^6^. **A:** Body-surface potential measurement and cardiothoracic imaging (CT or MRI) to visualize electrode positions and heart geometry. **B:** Recorded body-surface potentials before inverse reconstruction. **C:** Epicardial reconstructed potentials for each virtual node on the ventricular surface. **D:** By determination of activation time (AT, steepest downslope of local QRS complex in C) and recovery time (RT, steepest upslope of local T-wave^11^ in C), activation and recovery isochronal maps were calculated. **E:** Epicardial bullseye projection of isochronal AT and RT maps, with anatomical reference shown in the middle. **F:** Bullseye projection of activation gradients, activation-recovery interval (ARI) and recovery gradients, all derived from **D**.

### Data processing and ECGI reconstruction

For each subject, eleven normally-conducted sinus beats were selected from the body-surface recording (Figure 1B) for inverse reconstruction. Out of these eleven beats, ten were consecutive. One beat at a different point in time (separated by 268 (182-516) seconds, depending on recording duration) was randomly selected as a remote reference for comparison. For subject 5, a premature ventricular complex (PVC) was also selected for reconstruction. Our previously-validated ECGI methods (Supplementary Figure 1 and 9,^10^) were then used to reconstruct unipolar electrograms (UEG) from the body-surface potentials onto the ventricular epicardial surface (Figure 1C). For each epicardial UEG, the activation time (AT) and recovery time (RT) were automatically determined from the steepest downslope of the epicardial QRS complex and the steepest upslope of the epicardial T-wave^11^, respectively, using a spatiotemporal approach that considers the spatial flow of current^12^. Isochronal AT and RT maps were visualized on the 3D heart surface (Figure 1D). For more details, see Supplementary Methods.

### Standardized bullseye visualization

Our previously-validated open-source algorithm (UNISYS^13^) was used for standardized epicardial bullseye visualization of the results (Figure 1E), allowing segmental analysis and comparison between different hearts. Results were compared between left and right ventricles (LV and RV, respectively), circumferential segments (apex/center/base) and radial segments (LV-lateral/anterior/RV-lateral/inferior).

### Outcome measures

Metrics were determined from local AT and RT (Figure 1F). Activation-recovery interval (ARI), a surrogate measure of local action-potential duration^11^, was calculated as the subtraction of local AT from local RT. Duration of activation (AT_DUR_) and recovery (RT_DUR_) were quantified as the difference between the maximum and minimum AT and RT, respectively. Local spatial gradients of activation (ATG) and recovery (RTG) were defined as the largest local gradient (AT or RT difference divided by internode distance) in a 10-mm region. Activation-ARI relationships were investigated using linear regression with AT as the independent variable and ARI as the dependent variable (ARI = α ⋅ AT + β).

### Statistics

Unless otherwise noted, all outcome measures are averaged over 10 consecutive sinus beats to ensure consistency and minimize the influence of local outliers. Outcome measures are presented as average ± standard deviation (SD) if normally distributed, and median (first-third quartiles) are provided for non-normally distributed data (tested with the Shapiro-Wilk test). Unpaired two-tailed Student’s t-tests were used to compare normally distributed data, and the Mann-Whitney-U test was used to compare non-normally distributed data. P ≤0.05 was considered significant. Outcome measures were compared between males and females, and between subjects <50 years (y) and ≥50 y of age, resulting in two groups of approximately equal size. To compare the recovery of subjects <50 y with that of subjects

≥50 y, we selected those with similar RR intervals, as this influences recovery. Consequently, 7 subjects were selected for each age group, with comparable RR intervals.

Intra- and inter-individual Pearson’s correlation coefficients (CCs) were calculated to compare ECGs, AT maps and RT maps between and within subjects. For isochrone bullseye comparisons, we compared ATs and RTs at 20 standardized locations (the center of each bullseye segment, see Figure 1E). For between-subject comparisons, the inter-individual CC of the fourth beat is reported, to ensure a regular heart rate when feasible. For within-subject comparisons, we report the average value of all consecutive beat-to-beat CCs (beats 1 vs. 2, 2 vs. 3, etc.) and the average CC of all consecutive beats vs. the beat remote in time (beat 1 vs. remote beat, beat 2 vs. remote beat, etc).

## Results

### Characteristics of study subjects

**Table 1** summarizes the subject characteristics. Average age of normal subjects was 49±14 years and 6/22 were male. Average height of normal subjects was 179±6 cm for males and 167±6 cm for females. Median weight was 73 (62-80) kg for males and 65 (57-74) kg for females. Left ventricular ejection fraction, QRS duration and QTc according to Bazett’s formula were within the normal range in all individuals.

### Electrical activation and recovery for LV and RV

Figure 2 shows activation and recovery maps for a male and female subject of a single normally-conducted sinus beat, both showing earliest epicardial activation at the RV anterior base, with additional breakthroughs at other sites. Local UEGs at breakthrough sites showed an ‘rS’ morphology, indicative of an endo-to-epicardial activation pattern. Regional recovery patterns differed between these individuals.

**Figure 2:**
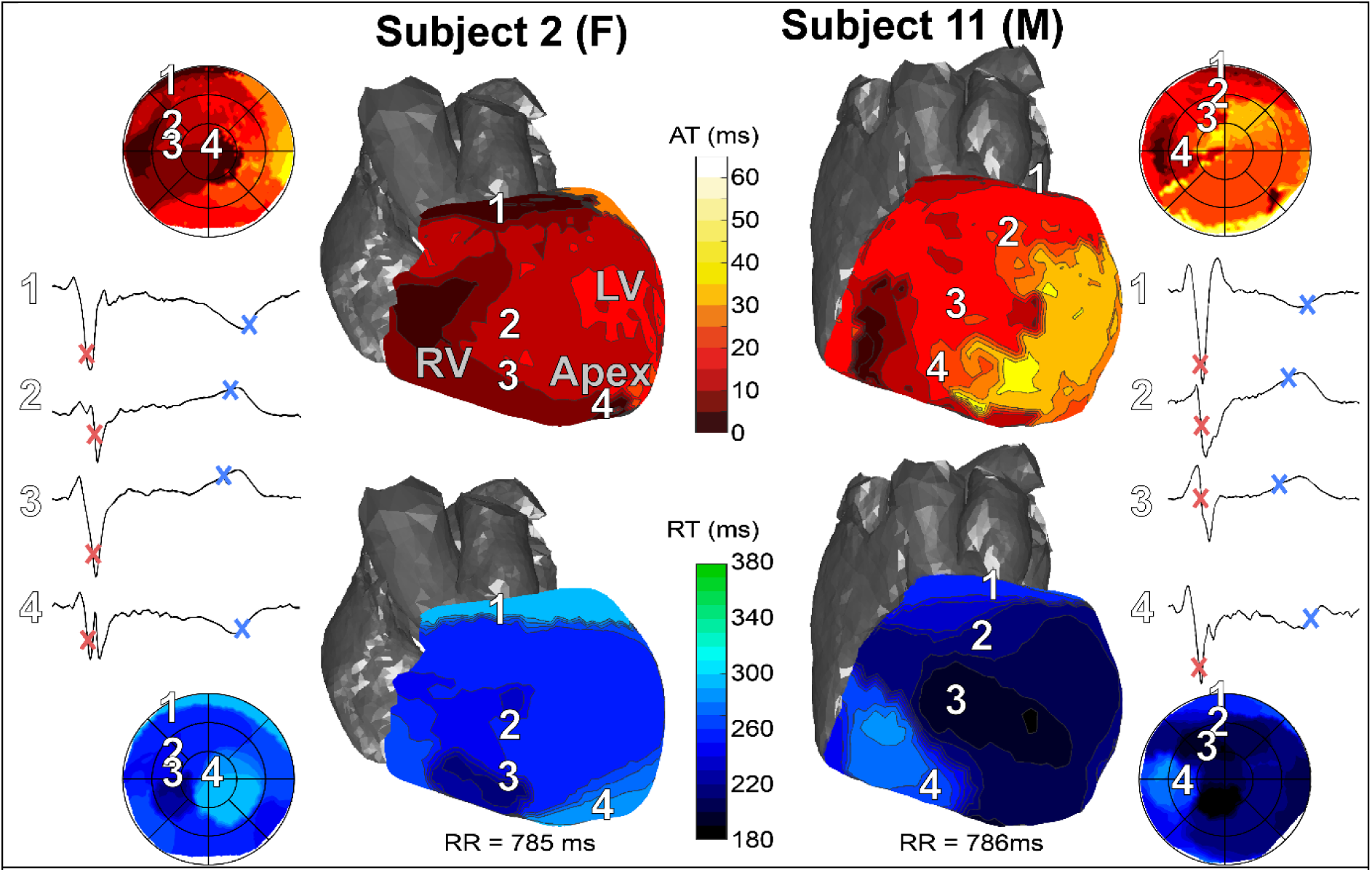
Illustrative examples of isochronal activation (top) and recovery (bottom) maps for a female (left) and male (right) subject. Numbers 1-4 indicate different locations on the heart and their corresponding electrograms. AT and RT are shown by red and blue markers, respectively, on these electrograms. Bullseyes in the corners of the figure show the entire epicardium whereas 3D views only show the anterior side.

Activation and recovery characteristics for all 22 normal subjects averaged for 10 consecutive beats are shown in Figure 3A-B. Average AT of the LV was 4 ms later than that of the RV (p<0.01). Average AT did not differ significantly between the sexes (p=0.4) or age groups (p=0.96).

**Figure 3:**
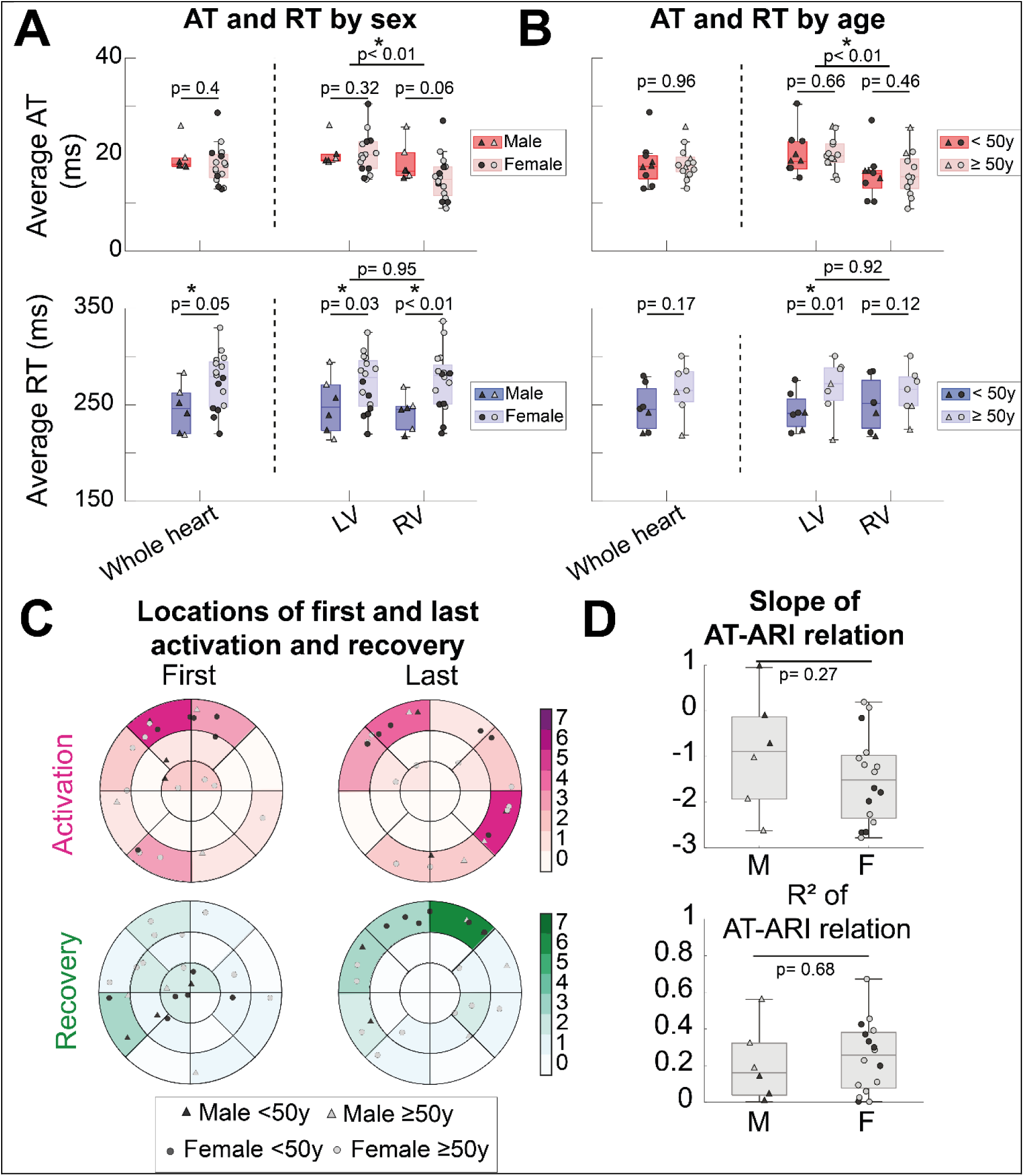
Overview of control results. A: Average AT and RT for males and females. The top horizontal bar indicates p-value between LV and RV. Second-level horizontal bars indicate p-values between males and females. Triangles indicate males, circles indicate females, filled symbols indicate subjects <50y and open symbols indicate subjects ≥50y. B: Average AT and RT for subjects <50y and ≥50y. (For RT, only 7 subjects were included for each group, to ensure similar RR intervals). C: Locations of first and last areas of activation and recovery. D: Slope and coefficient of determination of AT-ARI relation; see Results.

Average RT did not differ significantly between LV and RV (p=0.95) but was shorter in males than in females (246±25ms vs. 274±29ms, p=0.05). Recovery started significantly earlier in males (188±25 vs. 222±26; p=0.01) but duration of recovery tended to be longer, with a borderline difference (116 (114-120) vs. 96 (85-111) ms; p=0.06), see Supplementary Figure 2. RR intervals were not significantly different (904±184 ms in males vs. 905±132 ms in females, p=1.00). Average RT of LV was significantly shorter in subjects <50 y than in those ≥50 y (239±20 vs. 267±28 ms, respectively; p=0.01), but not for the RV or whole heart. RR intervals were not significantly different between these groups (828±107 ms vs. 875±81 ms, respectively; p=0.38).

Normalizing AT and AT_DUR_ for ventricular surface area did not result in significant differences between sex or age groups (p>0.26 in both cases). Additionally, AT and AT_DUR_ did not correlate significantly with ventricular surface area (p>0.25 in both cases). Further details on segmental analyses of AT and RT are provided in Supplementary Figure 3.

### Locations of first and last activation and recovery

Figure 3C shows the locations of the first and last activation and recovery of a normally- conducted sinus beat for all normal subjects. First epicardial activation appeared at the anterior/anterolateral RV in 10/22 subjects. The location of last activation occurred most often at the anterior/anterolateral RV (9/22) or inferior/inferolateral LV (8/22). Earliest recovery typically occurred on the RV (14/22), whereas last recovery was more heterogeneous between subjects. In subjects <50 y, the locations of first and last recovery appeared to be less dispersed than in subjects ≥50 y.

### Activation-ARI relationship

The normal T-wave on the 12-lead ECG is typically considered concordant with the QRS complex because of early-activated tissue having a longer ARI and later-activated tissue having a shorter ARI. Activation-ARI relationships (defined as ARI = α ⋅ AT + β) capture the relationship between local repolarization duration and the activation sequence. These had a negative slope on average, -1.30±1.08 (-0.88±1.3 for males vs. -1.46±0.99 for females; p=0.27), although slopes were heterogeneous between subjects, see Figure 3D. The average coefficient of determination (R^2^) was 0.24±0.19 (0.21±0.21 for males vs. -0.25±0.19 for females; p=0.68).

### Between-subject and within-subject variability

Figure 4 shows the comparison of ECG lead II and their co-registered activation and recovery bullseyes in all normal subjects. Whereas the ECG was normal, high-resolution AT and RT maps showed considerable variation among individuals. Figure 5A shows the ECGs of two subjects, together with their co-registered AT and RT maps. Within-subject AT and RT maps remained stable over 10 beats. 12-Lead ECGs, AT and RT maps were quantitatively compared within and between subjects using CCs, see Figure 5B. The CC for 12-lead ECG comparisons of the fourth beat between subjects was 0.61(0.46-0.72). CCs for between-subject comparisons of AT maps and RT maps were 0.32(0.07-0.50) and 0.20(-0.12-0.42), respectively. On the other hand, CCs of within-subject beat-to-beat comparisons of ECG, AT maps and RT maps were 0.98(0.97-0.99), 0.92(0.87-0.94) and 0.74(0.71-0.79), respectively, indicating a high within-subject stability of these outcome measures. When comparing the 10 consecutive beats within a subject to a beat remote in time, CCs were 0.95(0.93-0.97) for comparisons of the 12-lead ECG, 0.82(0.80-0.91) for AT maps and 0.78±0.08 for RT maps.

**Figure 4:**
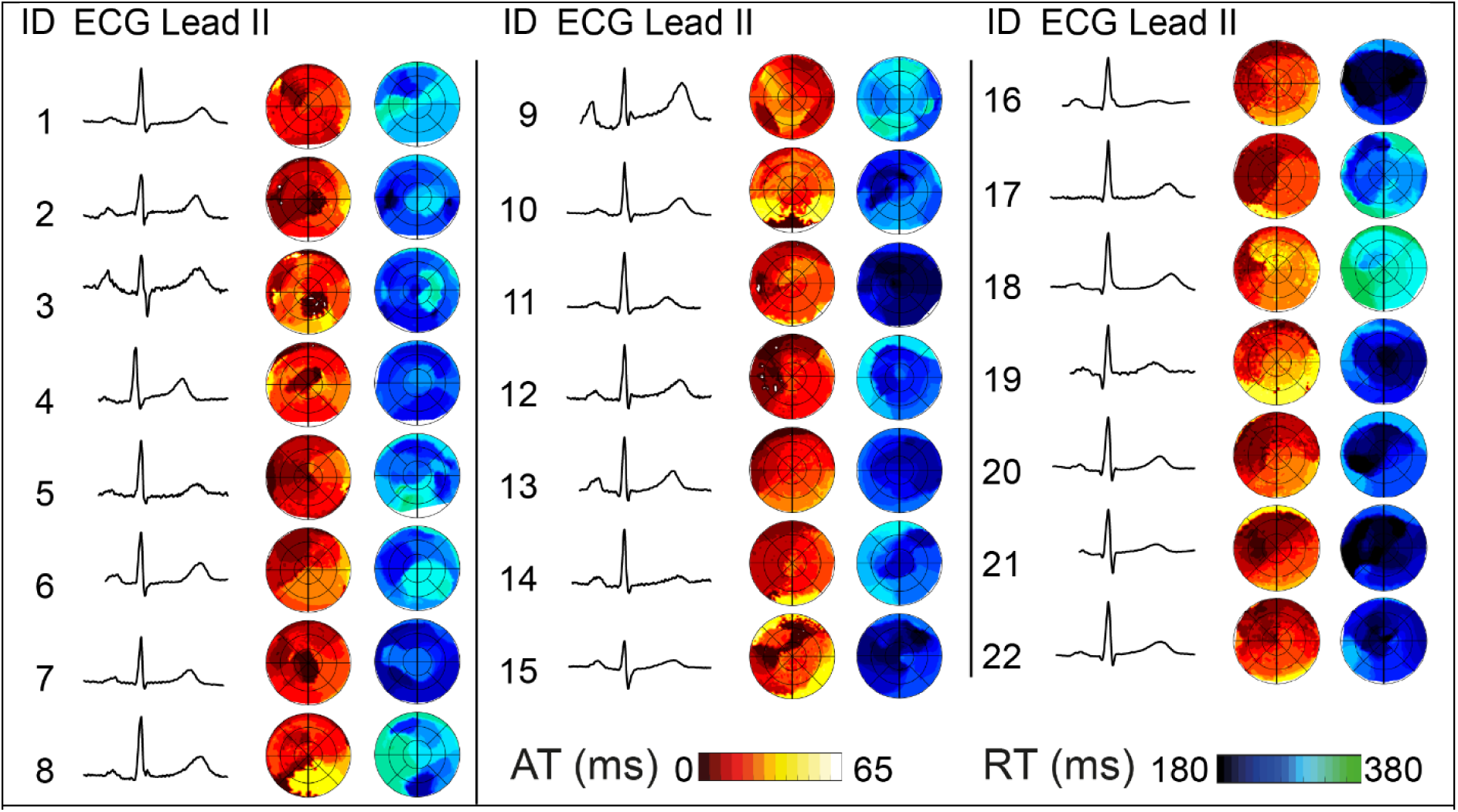
Lead II of the body-surface ECG, together with their co-registered activation isochrones and recovery isochrones for all subjects.

**Figure 5:**
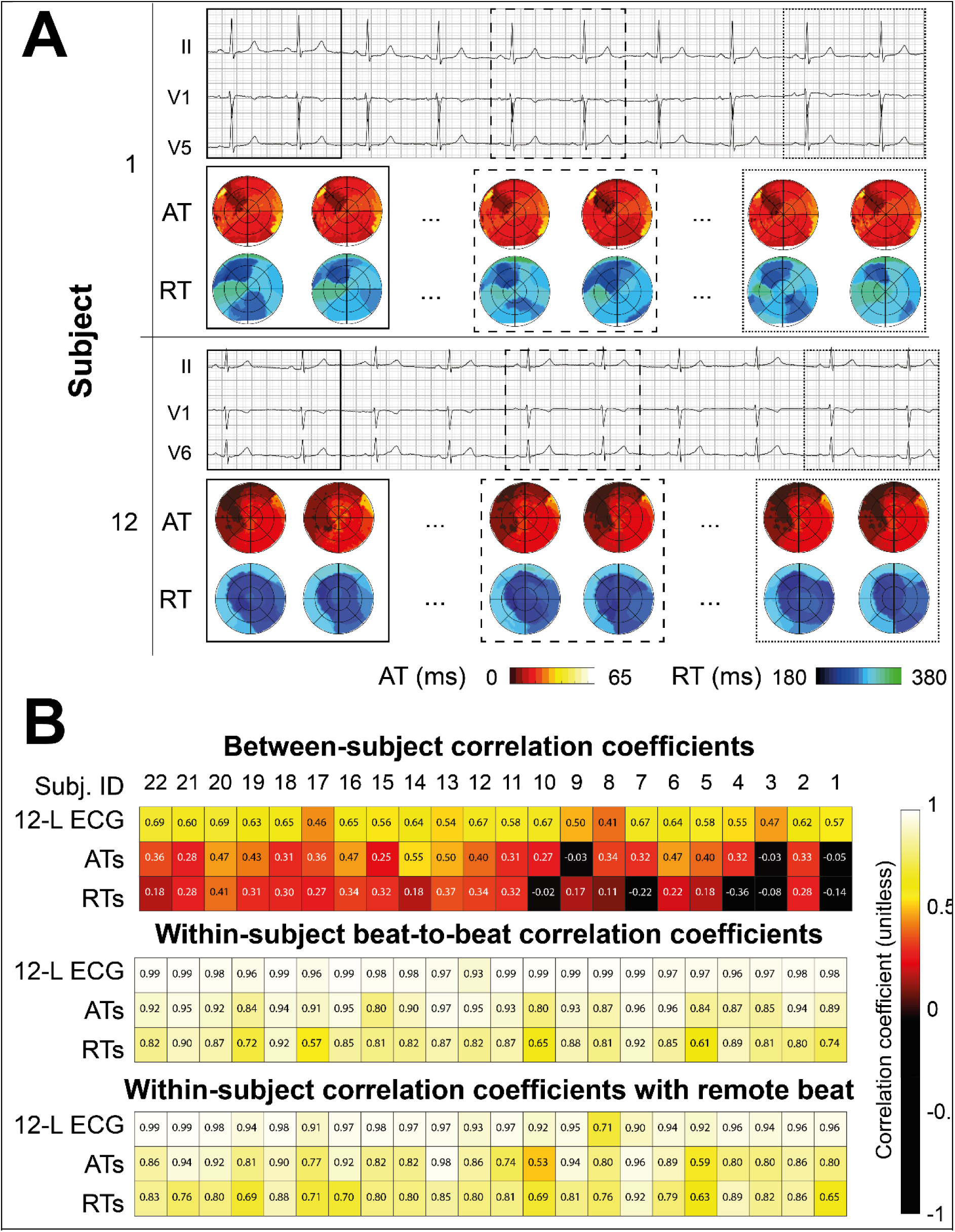
Within-subject and between-subject comparisons of ECG, activation maps and recovery maps. A: 3-lead ECG and corresponding isochronal activation- and recovery bullseyes of two selected subjects (1 and 12). For each subject, activation and recovery of beats 1, 2, 5, 6, 9 and 10 are shown. Square boxes indicate which bullseyes correspond to which beats. B: CCs are used to compare 12-lead ECGs, activation maps, and recovery maps between and within subjects. E.g., for between-subject comparisons, the median CC is noted for between-subject comparisons of subject 1 to subjects 2-22. For within-subject comparisons, the median value is noted for beat-to-beat comparisons of beat 1 vs. beat 2, beat 2 vs. beat 3, etc.

### Bundle-branch block

Figure 6 shows the activation and recovery patterns of a control subject, the RBBB subject, and the LBBB subject with and without biventricular pacing (BiV). In the control subject, the RV activated first, AT_DUR_ was 45 ms and RT_DUR_ 135 ms. A premature ventricular complex (PVC) from the inferolateral RV caused nonsynchronous activation, resulting in an AT_DUR_ of 83 ms and an RT_DUR_ of 95 ms.

**Figure 6:**
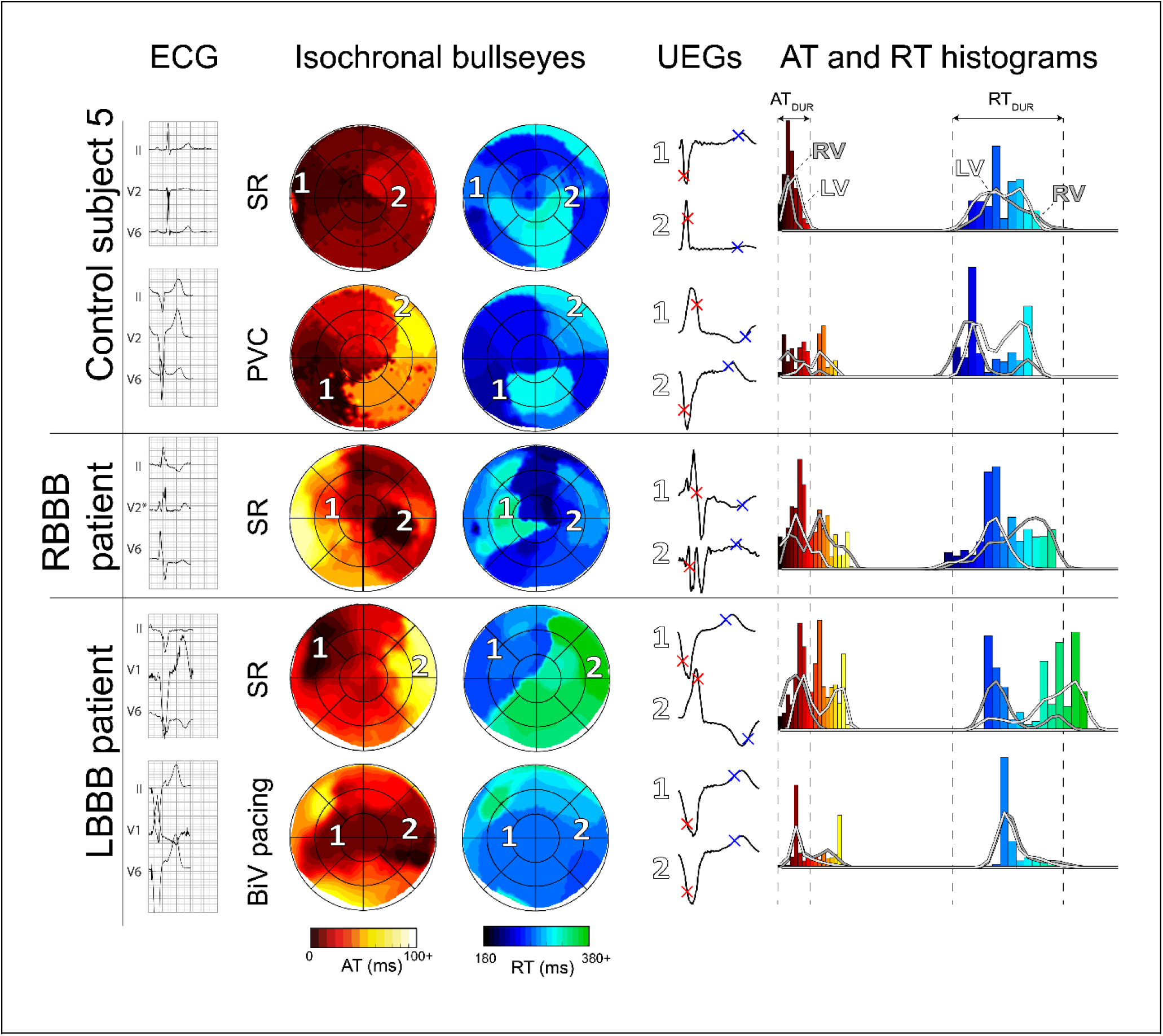
Comparison of activation and recovery patterns of a control subject in sinus rhythm (SR) and with a premature ventricular complex (PVC) (top), a subject with right bundle branch block (RBBB, middle) and a subject with left bundle branch block (LBBB) during SR and with biventricular (BiV) pacing (bottom). Body-surface ECGs, isochronal bullseyes with selected unipolar electrograms (UEGs) and AT and RT histograms are shown from left to right. (Dys)synchrony in activation and recovery between left and right ventricle (LV and RV), as visible in isochronal bullseyes, is also reflected in the histograms.

In the RBBB subject, the nonsynchronous activation caused the LV to activate first, resulting in a prolonged AT_DUR_ of 94ms and RT_DUR_ of 204ms. The LV activated and recovered before the RV.

In the LBBB subject, the nonsynchronous activation pattern caused an AT_DUR_ of 87 ms and RT_DUR_ of 126 ms. The RV activated and recovered before the LV. Short-term BiV of several beats resulted in comparable AT_DUR_ (91 ms) as during sinus rhythm, but partially resynchronized the LV and RV, resulting in early-activated areas in both ventricles and more homogeneous recovery (RT_DUR_ 99 ms). Results of all BBB subjects are further compared quantitatively with controls in Supplementary Figure 4. The agreement of our findings with literature data is shown in Supplementary Figures 5 (for activation-based measures) and 6 (for recovery-based measures).

### Long-QT syndrome

Figure 7 shows the comparison of activation and recovery patterns between a control subject and four subjects with congenital LQTS. The control subject, age- and sex-matched to the asymptomatic LQT3 patient (see Table 1), had an average AT of 13 ms and an average RT of 272 ms. Both symptomatic LQTS patients exhibited a prolonged QT interval and T-wave negativity, as seen on the ECG. This was also evident from ECGI, resulting in an average RT of 441 and 429 ms; average AT was 20 and 25 ms, within the normal range. The asymptomatic LQTS patients had a normal QT interval on the ECG and a borderline average AT (28 and 34 ms). However, ECGI revealed an average RT of 307 and 315 ms, markedly longer than age- and sex-matched controls. Quantitative comparisons of pathological subjects to age- and sex-matched controls are provided in Supplementary Figure 4. All values for ECGI-based metrics are provided in the Supplementary Data file for future reference.

**Figure 7:**
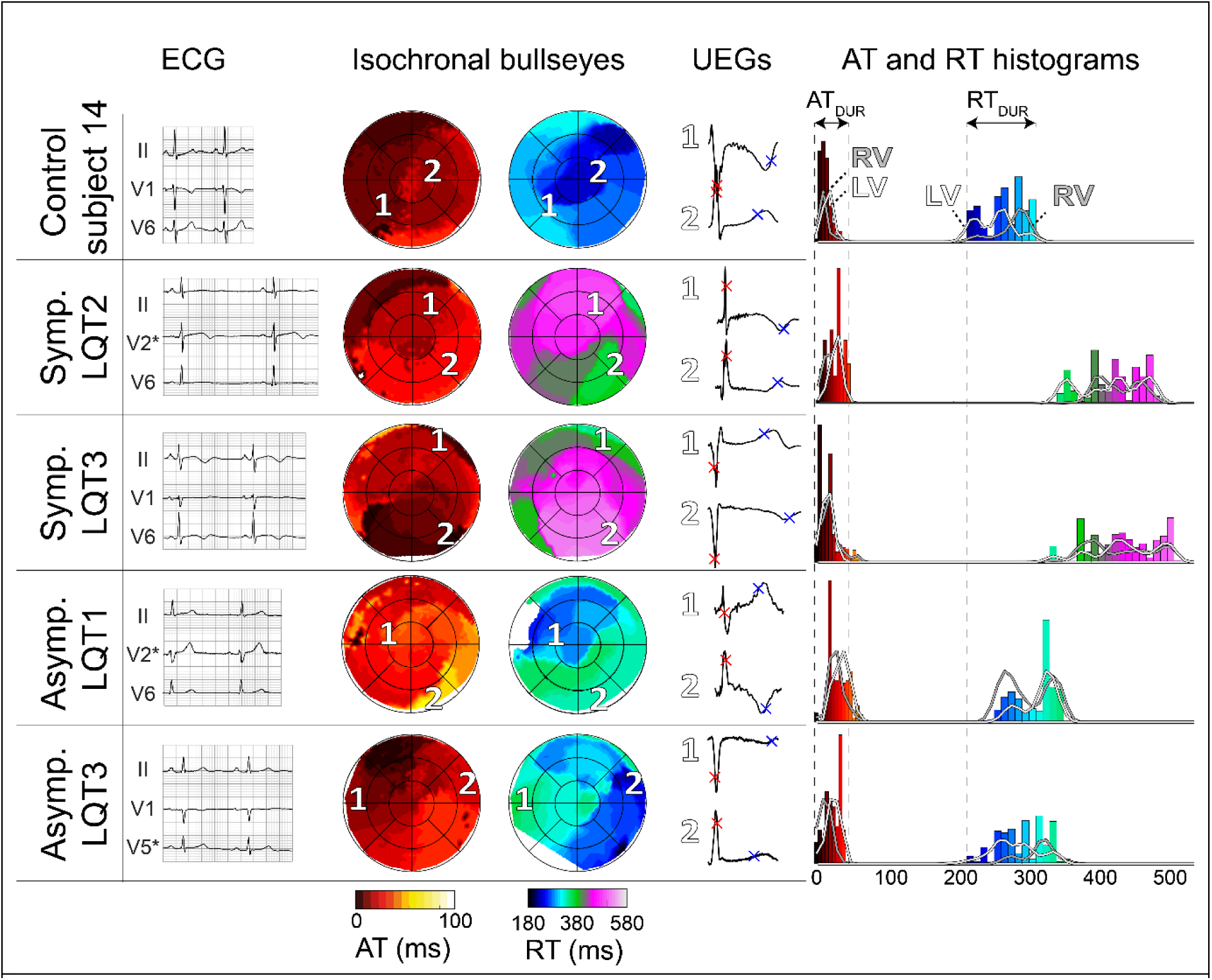
Comparison of activation and recovery patterns of five subjects in sinus rhythm: a control subject (top), two clinically-overt LQTS patients with history of ventricular tachyarrhythmias (“Symp. LQT2” and “Symp. LQT3”) and two clinically-occults without such history (“Asymp. LQT1” and “Asymp. LQT3”). Body-surface ECGs, isochronal bullseyes with selected unipolar electrograms (UEGs) and AT and RT histograms are shown from left to right. (Dys)synchrony in activation and recovery between left and right ventricle (LV and RV), as visible in isochronal bullseyes, is also reflected in the histograms.

Agreement of our study results with literature data is shown in Supplementary Figure 6.

## Discussion

Using noninvasive ECGI, we have demonstrated that ventricular activation and recovery patterns can vary considerably among normal individuals, who all had unremarkable 12-lead ECGs. In each of these subjects, activation and recovery characteristics were relatively stable over the course of multiple beats to minutes. Recovery times were shorter in males than in females, and at younger age (in the LV). We found only limited association between the sequence of activation and the duration of local recovery (ARI).

### Activation and recovery patterns vary greatly between control subjects, but are stable within individuals

Although all subjects had a normal 12-lead ECG, their underlying ventricular activation and recovery patterns were profoundly different. This appears consistent with the results of previous studies, although differences were not reported explicitly there^4,5^. Individual characteristics such as conduction system anatomy, Purkinje network distribution, ion-channel expression and autonomic innervation of the ventricular myocardium may underlie the variant activation and recovery patterns that we have revealed, and they may exaggerate under pathological conditions. The relative stability over multiple sinus beats in individual study subjects implicates that electrical perturbations during pathological conditions may be signalled by ECGI by revealing abnormal dynamics in the activation and recovery isochrones, potentially even before the emergence of ectopy, as is also hinted by recent other work^14^. Likewise, dynamic alterations in the substrate, e.g., in relation to arrhythmic events or during drug provocation testing, could be uncovered.

### Activation characteristics

The epicardial locations of first (anterior to anterolateral RV) and last (RV anterior base or LV lateral base) activation that we found are consistent with findings from earlier studies^5,7,8,15,16^. Interventricular variations in wall thickness, myofiber orientation and ion-channel expression^4^ likely contributed to the sites of first and last activation, and to the shorter average RV AT compared to the LV. Activation duration was similar to previous ex-vivo^5^ and in-vivo^15^ studies and 8 ms longer than in a noninvasive mapping study^7^.

No significant differences were found in average AT or AT_DUR_ between males and females. Most activation characteristics were similar between the age groups (<50y vs. ≥50y), similar to a recent noninvasive mapping study ^17^.

### Recovery characteristics

Epicardial locations of first and last recovery were more variable between subjects than those of activation. First recovery mostly occurred in the RV anterobasal segments, whereas last recovery mostly occurred in RV/LV anterobasal segments. This is in agreement with findings in three explanted human hearts^4^. Epicardial recovery duration was similar to previous experimental work^4^ and ARIs (246 (219-264) ms) were similar to an earlier noninvasive mapping study^7^. There was a higher inter-individual variation of recovery than that of activation. Within-subject correlations of beat-to-beat RT maps were also lower than those of AT. RT may thus be more variable between beats, but we cannot rule out that the larger variation derives, at least partly, from computational difficulties of determining RT^18^.

No significant differences were found in average RT or RT_DUR_ between LV and RV. This contrasts with the earlier study of three explanted hearts^4^ but is in line with an earlier noninvasive mapping study^7^.

Males typically had a shorter average RT than females, as male electrical recovery started earlier and was not compensated by a longer recovery duration (higher value of [last RT – first RT]). Similarly, we found that average RT was shorter in younger (<50 y) than older subjects (≥50 y), as shown before ^17^.

### Activation-ARI relationship

Previous human studies investigating the activation-ARI relationship were performed in diseased patients^1,2,19^ or ex-vivo hearts^3^, but in-vivo results in normal subjects under physiological conditions were lacking. We found no strong relationship between the epicardial activation timing and ARI. The AT-ARI relationships over the epicardium (LV *plus* RV) were negative but with weak correlation (slope -1.30±1.14 and R^2^ 0.23 (0.07-0.35)). This indicates that the local ARI does not only depend on activation, but probably involves many other properties such as electrotonic coupling, local ion-channel expression, autonomic innervation, and hormonal influences. These results contrast with previous in-vivo studies^1,2^, but align with an ex-vivo study ^3^, which may be explained by differences in study cohorts, mapping techniques, settings in which the recordings were performed, and the extensiveness of mapping.

### Pathological conditions

Activation (and consequently, recovery) patterns in subjects with BBB were different from controls. LBBB and RBBB resulted in less synchronized and prolonged activation, in agreement with previous ECGI findings^16,20–22^. Subsequent biventricular pacing for LBBB resulted in an increased LV-RV synchrony of activation and recovery in the LBBB subject. While increased activation synchrony is thought to be supportive of mechanical function, increased recovery synchrony is considered antiarrhythmic^10^. Electrical recovery was significantly prolonged in the subject with clinically-overt LQTS (patient A) in both the 12- lead ECG and ECGI, in agreement with previous ECGI findings^23^. Moreover, the RT histogram was more widespread, indicating dispersion. Whereas the 12-lead ECG of the subject with clinically-occult LQTS (patient B) was normal, ECGI revealed prolonged recovery compared to age- and sex-matched controls, which would not have been recognized otherwise.

Consistent with other literature reports, our results illustrate the value of ECGI in noninvasively detecting and localizing AT and RT abnormalities with high detail, thus enabling a better characterization of proarrhythmic substrates, the dynamics of arrhythmia and the effects of therapy.

### Study limitations

We used our previously-validated implementation of ECGI^9,24^ to noninvasively assess epicardial electrophysiology, which did not allow to directly study endocardial activation and recovery. In some aspects, noninvasive mapping is less accurate than invasive mapping^6,9^.

However, invasive mapping is an expensive, complication-sensitive and time-consuming method, and it would be unethical to perform this in non-diseased subjects. ECGI (with ∼1- cm resolution) does not reach the level of detail of invasive mapping (with presumed submillimeter resolution), but still enables to vastly enhance our understanding beyond the 12-lead ECG (without challenging the undisputed clinical value of the ECG). For a detailed overview of ECGI’s accuracy and a dissemination of its limitations, we refer to ^6^.

Our standardized bullseye visualization (UNISYS^13^) enabled standardized comparisons between epicardial segments within and between subjects, but thereby omitted the location of 3D anatomic structures that may vary between subjects, such as the outflow tracts and coronary arteries. Although we distinguished between males and females, and young and old subjects, we did not account for other potential confounding factors, such as the menstrual cycle phase, autonomic tone or time of recording (all of which could have influenced activation and recovery duration, RR interval, ion-channel expression and/or arrhythmia susceptibility). Finally, our cohort was predominantly female. Although women have been historically underrepresented in cardiovascular research, we recognize the possibility that our results could be biased by the relative weight of female characteristics.

## Conclusion

Despite similar 12-lead ECGs, electrical activation and recovery vary significantly between normal individuals, while remaining stable within the same individual over multiple consecutive beats and over minutes of time. Our study sets a new standard for understanding ventricular electrophysiology in normal males and females of different ages. Individual ECGI characteristics may serve as a basis to better understand and monitor the development of electropathologies and arrhythmia.

## What’s new?

- Despite similar unremarkable 12-lead ECGs, ventricular electrical activation and recovery patterns vary significantly between normal individuals. These differences potentially account for variations in the individual susceptibility to arrhythmias.
- Ventricular electrical activation and recovery are relatively stable over consecutive beats and over several minutes under normal conditions. Dynamic alterations revealed by ECGI may be causal or consequent to arrhythmic perturbations, and owing to the onset/progression of (genetic) cardiac disease.
- Our detailed results of ventricular activation and recovery sequences under physiological conditions provide a basis for better understanding of pathology. In future studies, we aim to investigate the relationship of these individual electrical characteristics at baseline with arrhythmia propensity.

## Data Availability

All data produced in the present study are available upon reasonable request to the authors

## Notes

### Competing Interest Statement

M.J.M.C. is part-time employed by Philips Research.

### Funding Statement

This work was supported by the Special Research Fund (BOF) of Hasselt University and Maastricht University Medical Center (MUMC+) [BOF17DOCMA15 to J.S.]; the Netherlands CardioVascular Research Initiative [CVON2017-13 VIGILANCE to J.S., B.v.R. and P.G.A.V., and CVON2018B030 PREDICT2 to P.G.A.V.]; a Kootstra Talent Fellowship research grant from Maastricht University [2015T61 to U.C.N.]; the German Academic Scholarship Foundation and the Walter Benjamin Programme by the German Research Foundation [DFG, 529532291 to P.M.D.]; the Dutch Heart Foundation [2021T016 to U.C.N.]; The Netherlands Organization for Scientific Research [(ZonMw (0915016181013)) to R.t.B. and TTW16772 to M.J.M.C.]; and the Health Foundation Limburg (Maastricht, The Netherlands) [to M.J.M.C].

### Author Declarations

Ethics committee of Maastricht University Medical Center+ (MUMC+), The Netherlands gave ethical approval for this work (METC 11-2-043). Ethics committee of Health Research Authority London, United Kingdom (Surrey; 19/LO/0762) gave ethical approval for this work

