## Supplementary material for "Variant electrical activation and recovery in normal human hearts revealed by noninvasive electrocardiographic imaging"

### Supplementary Methods

For a stable and accurate inverse solution, ECGI requires several key components: validation, processing of anatomical images, preprocessing of body-surface potential maps, and the inverse reconstruction. The inverse reconstruction can be further subdivided into the choice of ECGI formulation, the transfer matrix, regularization, and activation/recovery time determination and postprocessing. We provide a concise overview of methods here, for each of these components.

#### Validation

The ECGI methodology and data processing of the current study were similar to our previous validation studies ^1,2^. A summary of our in-vivo validation study can be found in Supplementary Figure 1.

#### Anatomical images

Electrode positions and epicardial geometry were manually segmented from the anatomical images, with the Seg3D software ^3^. For each individual, a torso geometry consisting of the centroids of electrode positions and a ~2000-nodal epicardial geometry were then digitized. The latter was generated by downsampling the exported segmentation and performing Laplacian smoothing in FEBio Preview ^4^.

#### Body-surface potential maps

Noisy body-surface signals were semi-automatically selected and removed, resulting in a remaining 173±30 electrode signals. Baseline drift and 50 Hz noise were removed from the remaining body-surface potentials. Individual beats were manually selected, and their QRS complexes and T-waves were manually annotated. T-waves were filtered with a second-order 40 Hz low pass Butterworth filter ^5^.

#### Inverse reconstruction

##### ECGI formulation

Previously validated ECGI methods ^1,2^ were used to reconstruct unipolar electrograms (UEG) on the ventricular epicardial surface for selected beats (Figure 1C). Inverse reconstruction of epicardial potentials was performed with the potential-based formulation of ECGI, which relies on the numerical relation between electrical potentials at the heart and body surface ^6^.

##### Transfer matrix

For each subject, we established the electrostatic relationship between the torso electrodes and epicardial ventricular nodes using a transfer matrix ^7^ through publicly available methods^8^. These methods assume homogeneous conductivity throughout the torso, resulting in comparable ECGI accuracy to an inhomogeneous torso model^9^, with improved ease of use. The transfer matrix is created with both the torso and heart considered non-moving. In-vivo and computational studies have shown that the effects of contractile and respiratory motion on common outcome measures in ECGI are small in physiological scenarios ^10,11^.

##### Regularization

Subsequently, we applied zeroth-order Tikhonov regularization ^12^ to constrain the solution to the inverse problem. For each cluster of either one or ten beats, a single Tikhonov parameter (λ) was used for all time instants to improve the temporal consistency of the reconstruction, which is important for UEG quality. This λ was chosen as the median of all values obtained with the L-curve method ^13^ for the different time instants during the QRS complexes and T-waves.

##### AT and RT determination

For each epicardial UEG, the activation time (AT) and recovery time (RT) were automatically determined from the steepest downslope of the epicardial QRS complex and the steepest upslope of the epicardial T-wave ^14^, respectively, using a spatiotemporal approach. This approach considers the spatial flow of current and is more accurate than a temporal-only approach ^15^. ATs and RTs were determined relative to the average moment of steepest QRS downslope of the first 25 activated nodes as a common time reference. Isochronal AT and RT maps were visualized on the 3D epicardial surface (Figure 1D).

##### RT postprocessing

To ensure correct annotation of RTs, 1) local RTs outside the 2^nd^-to-98^th^ percentile of all RTs were blanked, and 2) for each electrogram, RT was blanked if it fell outside the area of the largest positive derivative of the local T-wave, an adapted version of the previously published “confidence score” ^16^. A 15-mm spatial median filter was applied to remove noisy outliers and interpolate regions that were blanked ^2^. RTs were then manually verified. Finally, any areas of RTs that were manually determined to be erroneous (due to flat or noisy UEG T-waves) were blanked and spatially interpolated.

### Supplementary Figures

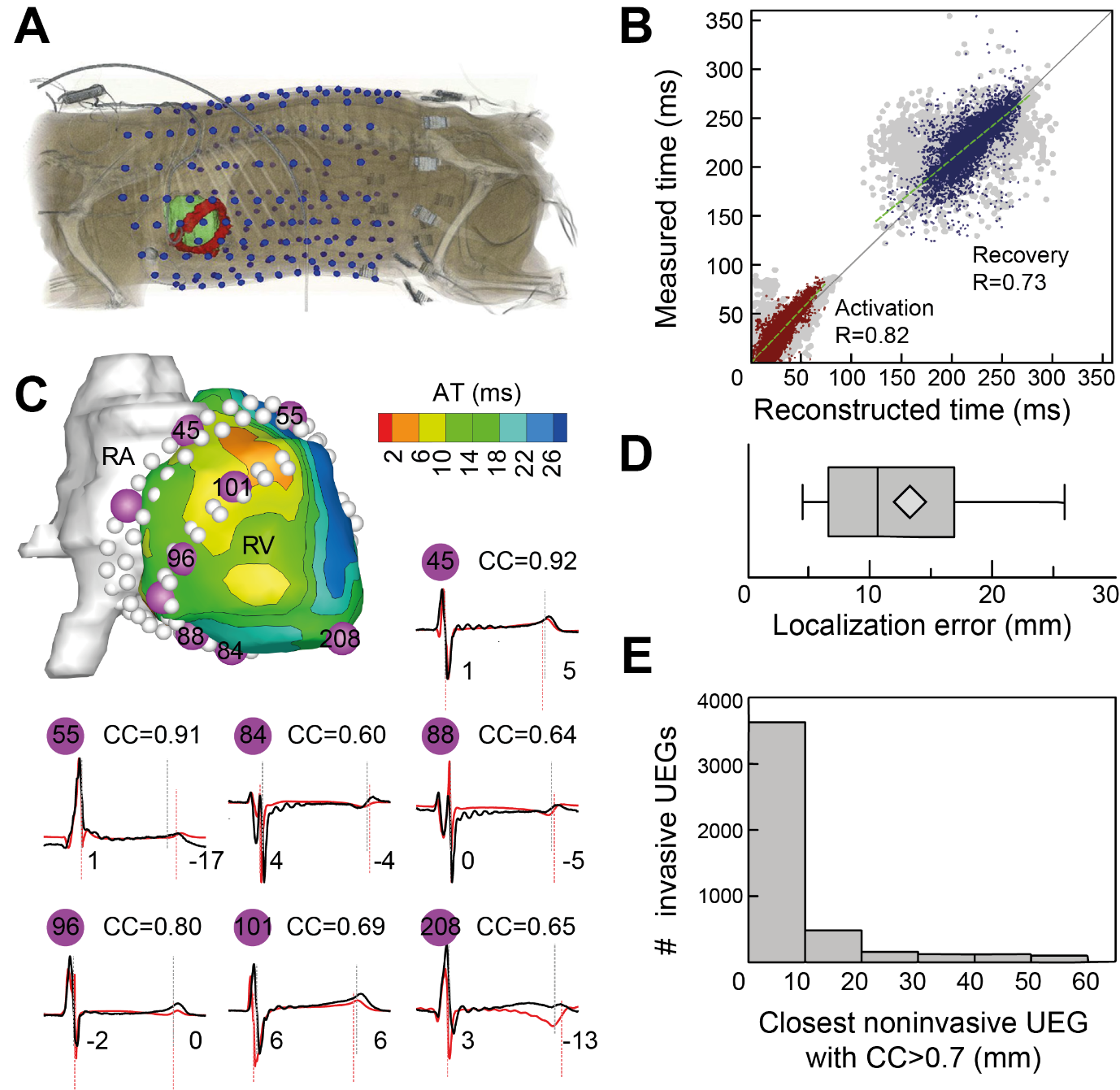

**Supplementary Figure 1:** In-vivo validation of ECGI^1^. **A:** Experimental setup as applied in normal anesthetized dogs, illustrating body-surface electrodes (blue), epicardial surface (green), and epicardial contact electrodes (red). **B:** Scatter plot of invasively-measured vs. ECGI-reconstructed activation (R=0.82) and recovery times (R=0.73). **C:** Epicardial surface, with purple spheres indicating the position of epicardial electrodes. For the numbered purple spheres, recorded (red) and ECGI-reconstructed (black) UEGs are depicted below. Grey and red dashed lines indicate recorded and reconstructed activation/recovery times, respectively. The mismatch between both is given in milliseconds at the right for each pair. **D:** Box plots for localization mismatch for 80 paced beats in 4 dogs. Localization mismatch is deﬁned as the distance between the known pacing location and the location of earliest activation from non-invasively reconstructed UEGs. **E**: Histogram showing the spatial accuracy of ECGI, defined as the distance of each invasive UEG to the nearest noninvasive UEG with a good-enough correlation (CC>0.7).

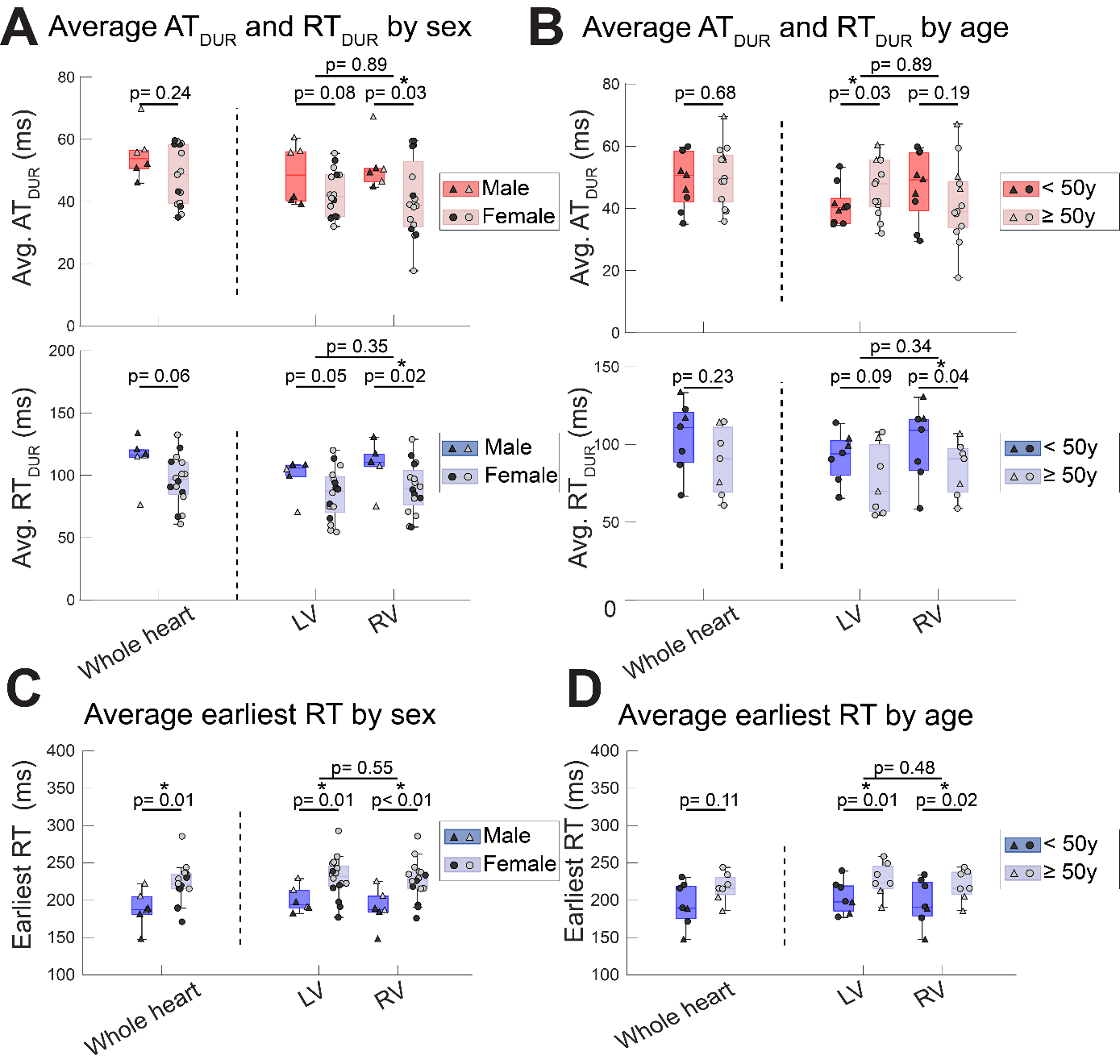

**Supplementary Figure 2**: Average activation and recovery duration (AT_DUR_ and RT_DUR_, respectively) by sex and age (<50 y vs. ≥50 y). The top horizontal bars indicate p-value between segments. Second-level horizontal bars indicate p-values between males and females. For RT comparisons in B and D, only 7 subjects were included for each group, to ensure similar RR intervals.

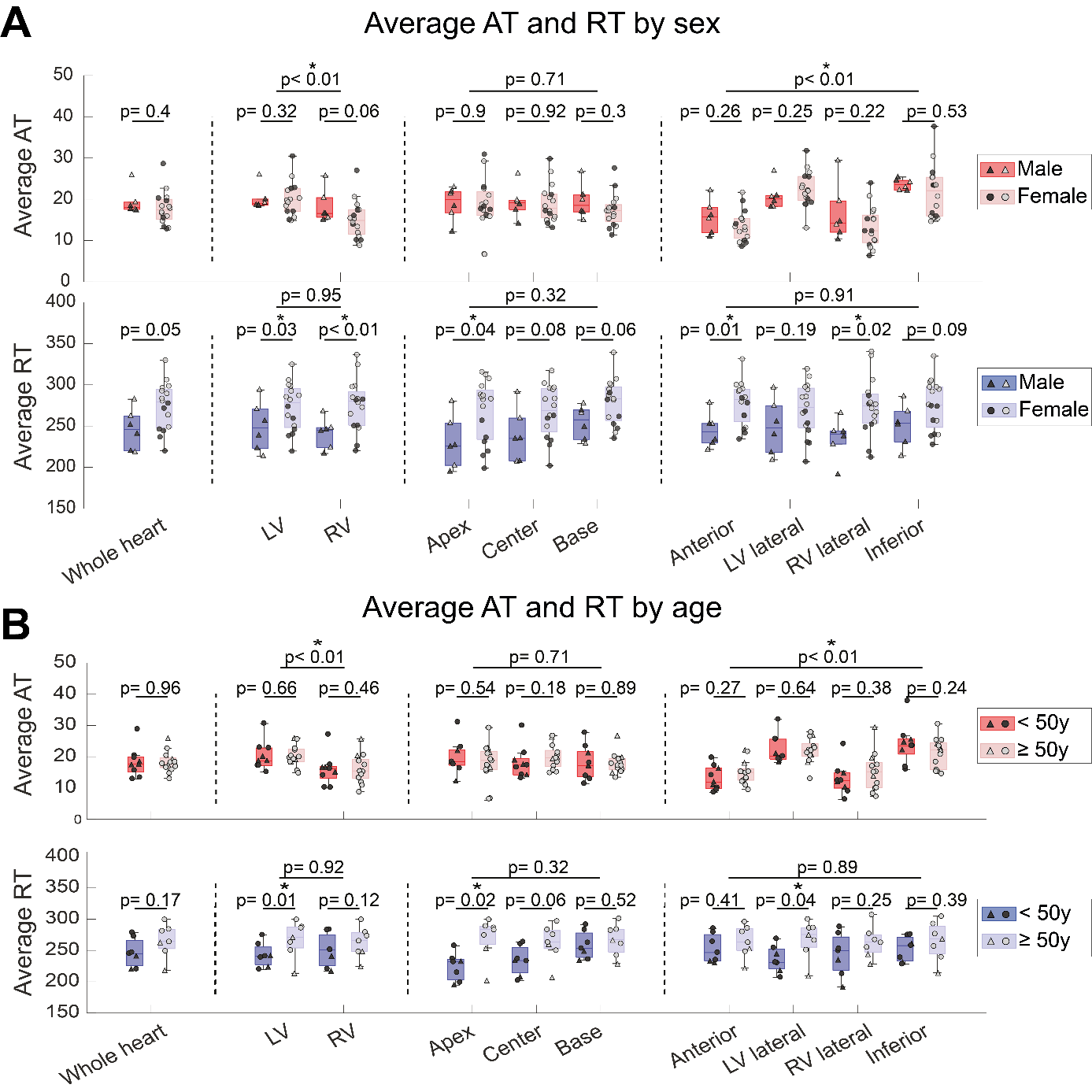

**Supplementary Figure 3**: Average AT and RT by sex (A) and age (<50 y vs. ≥50 y, B), for all segments. The top horizontal bars indicate p-values between segments. Second-level horizontal bars indicate p-values between males and females. For RT comparisons in B, only 7 subjects were included for each group, to ensure similar RR intervals.

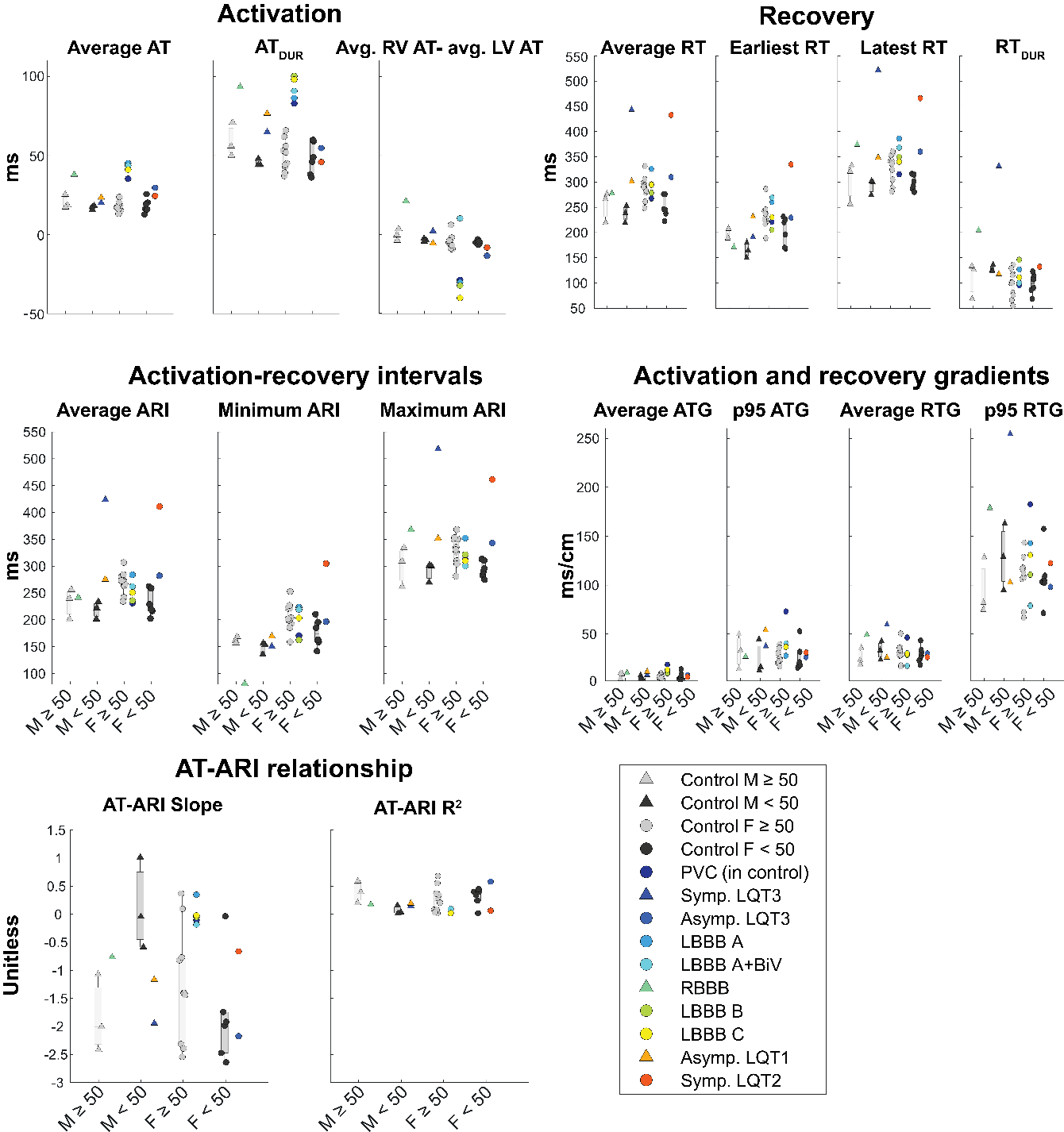

**Supplementary Figure 4:** comparison of important outcome measures between healthy and pathological individuals, stratified by age and sex. P95: 95^th^ percentile.

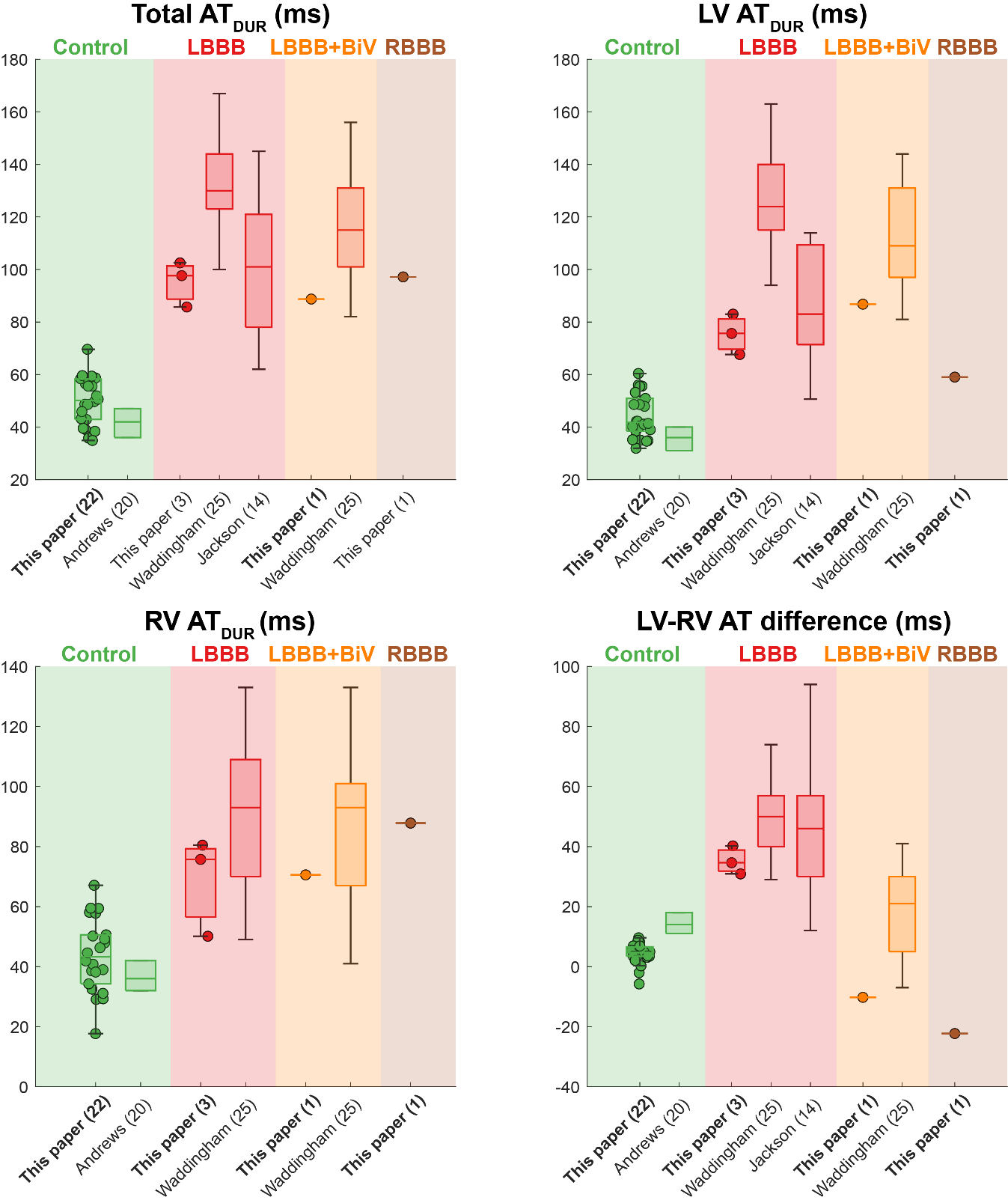

**Supplementary Figure 5:** Boxplots showing agreement of our activation-based study results with literature data^17–19^. Each graph indicates one outcome measure. Colors indicate conditions: control (green), LBBB (red), LBBB+BiV (orange), RBBB (brown). Within one color, different studies are shown on the x-coordinates, with the number of subjects shown in round brackets (*n*). Small circles indicate individual datapoints.

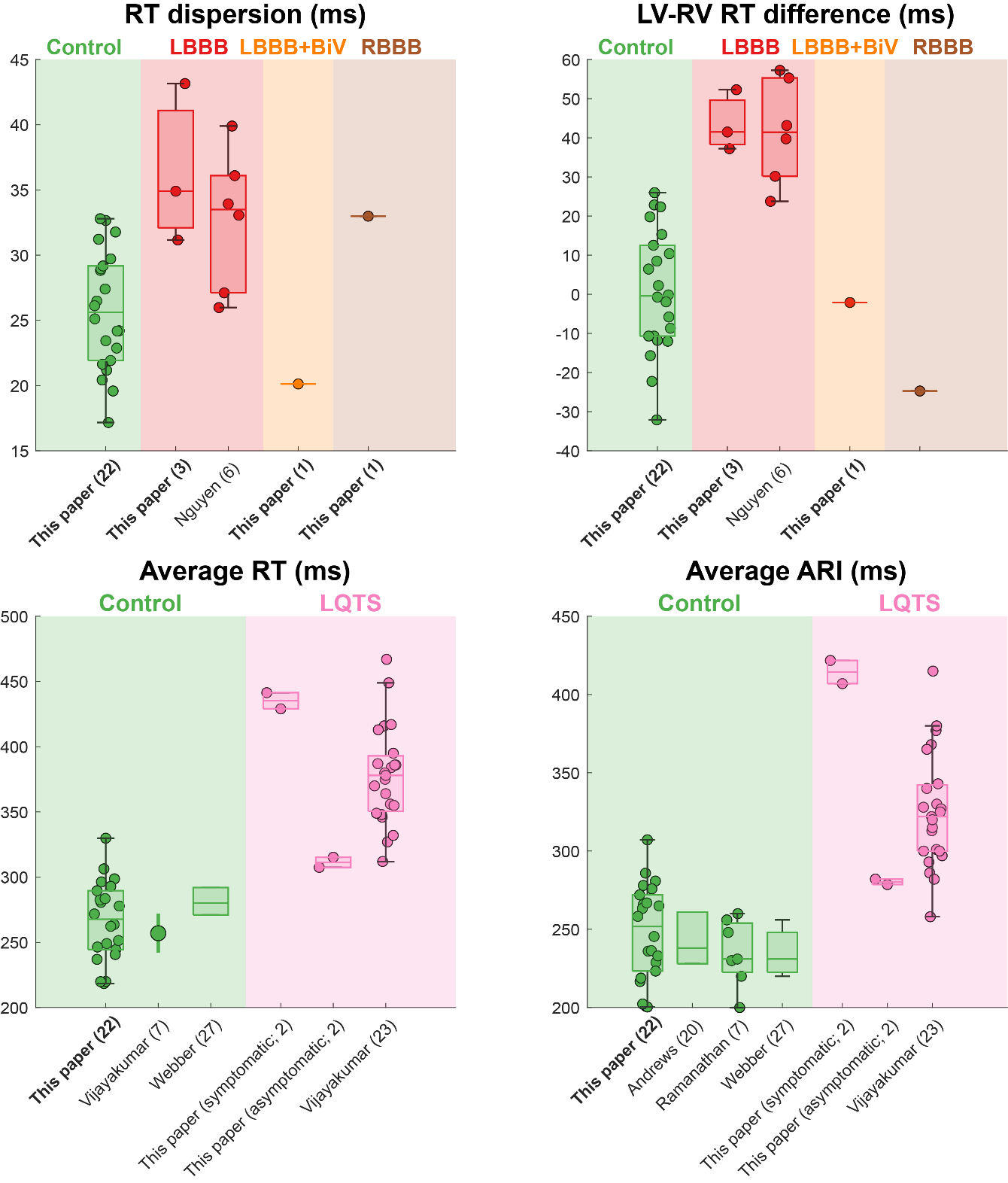

**Supplementary Figure 6:** Boxplots showing agreement of our recovery-based study results with literature data^17,20–23^. Each graph indicates one outcome measure. Colors indicate conditions: control (green), LBBB (red), LBBB+BiV (orange), RBBB (brown), LQTS (pink). Within one color, different studies are shown on the x-coordinates, with the number of subjects shown in round brackets (*n*). Small circles indicate individual datapoints, while large circles with vertical lines indicate average ± SD.
